# Is neglect of self-clearance biassing TB vaccine impact estimates?

**DOI:** 10.1101/2023.04.11.23288400

**Authors:** Danny Scarponi, Rebecca A Clark, Chathika Weerasuriya, Jon C Emery, Rein MGJ Houben, Richard G White, Nicky McCreesh

## Abstract

**Background:** Mathematical modelling has been used extensively to estimate the potential impact of new tuberculosis vaccines, with the majority of existing models assuming that individuals with Mycobacterium tuberculosis (Mtb) infection remain at lifelong risk of tuberculosis disease. Recent research provides evidence that self-clearance of Mtb infection may be common, which may affect the potential impact of new vaccines that only take in infected or uninfected individuals. We explored how the inclusion of self-clearance in models of tuberculosis affects the estimates of vaccine impact in China and India.

**Methods:** For both countries, we calibrated a tuberculosis model to a scenario without self-clearance and to various scenarios with self-clearance. To account for the current uncertainty in self-clearance properties, we varied the rate of self-clearance, and the level of protection against reinfection in self-cleared individuals. We introduced potential new vaccines in 2025, exploring vaccines that work in uninfected or infected individuals only, or that are effective regardless of infection status, and modelling scenarios with different levels of vaccine efficacy in self-cleared individuals. We then estimated the relative incidence reduction in 2050 for each vaccine compared to the no vaccination scenario.

**Findings:** The inclusion of self-clearance increased the estimated relative reductions in incidence in 2050 for vaccines effective only in uninfected individuals, by a maximum of 12% in China and 8% in India. The inclusion of self-clearance increased the estimated impact of vaccines only effective in infected individuals in some scenarios and decreased it in others, by a maximum of 14% in China and 15% in India. As would be expected, the inclusion of self-clearance had minimal impact on estimated reductions in incidence for vaccines that work regardless of infection status.

**Interpretations:** Our work suggests that the neglect of self-clearance in mathematical models of tuberculosis vaccines does not result in substantially biased estimates of tuberculosis vaccine impact. It may, however, mean that we are slightly underestimating the relative advantages of vaccines that work in uninfected individuals only compared to those that work in infected individuals.

## 1. Introduction

Tuberculosis is one of the leading causes of death from a single infectious agent worldwide, with an estimated 1.6 million deaths in 2021 [1]. The majority of countries are not on track to meet the WHO End TB Targets for 2035, which include reductions in the absolute number of tuberculosis deaths by 95% and the tuberculosis incidence rate by 90%, compared to 2015 levels. Modelling studies suggest that current available interventions are insufficient and that the development of new tools is essential to meet the WHO End TB Targets. The only licensed vaccine against tuberculosis, Bacillle Calmette-Guerin (BCG), is effective at preventing disseminated disease in infants, but most studies suggest that it offers limited protection against infectious pulmonary tuberculosis in adults [2]. The development of new tuberculosis vaccines, that protect adults from developing infectious disease, is therefore critical for reducing tuberculosis incidence and mortality in line with the End TB Targets [3].

There are currently several tuberculosis vaccine candidates in the clinical development pipeline, with the potential for diverse vaccine characteristics and indications. A 3-year phase 2b trial of the M72/AS01E candidate vaccine demonstrated an efficacy of 49.7% (95% confidence interval: 2.1–74.2) for preventing disease in adults who were positive by interferon-gamma release assay at baseline [4]. A trial of BCG-revaccination achieved an efficacy of 45.4% (6.4%–68.1%) against the secondary endpoint, preventing sustained infection, in a cohort of IGRA negative adolescents in South Africa [5].

Mathematical models have been used extensively to estimate the potential impact and cost-effectiveness of tuberculosis vaccines with different specifications [6]. Models have, for example, provided age indications for TB vaccines, by demonstrating the greater and more rapid impact before 2050 of targeting adolescents and adults instead of infants in low and middle income countries [7], [8], and older adults only in settings such as China [9]. Models have also assessed whether greater population-level impact would be delivered by vaccines with efficacy in Mycobacterium tuberculosis (Mtb)–infected populations (often referred to as current infection vaccines) versus uninfected populations (often referred to as no current infection vaccines) [7], [8], [10]–[12], and how population-level impact may vary by age and setting [10]. Model results such as these are being used to guide the development and implementation planning of new vaccines, informing, for example, the design of future clinical trials [7], [8], [13].

Most existing models of tuberculosis assume that individuals with Mtb infection remain at lifelong risk of progression to tuberculosis disease. However, recent research provides evidence that self-clearance of Mtb infection can occur: individuals previously infected with Mtb clearing their infections and ceasing to be at risk of progressing to TB disease (in the absence of reinfection) [14]. This is likely to have a substantial impact on tuberculosis epidemiology [15]. Due to self-clearance, the population with a viable Mtb infection may be markedly smaller than generally assumed, with such individuals at greater risk of progressing to TB disease. Since the efficacy of new tuberculosis vaccines may depend on the host infection status, self-clearance also has the potential to affect the impact of new vaccines. While a number of more recent vaccine models have included a simple representation of self-clearance ([6], [8]), none have investigated its effects on vaccine impact, or have explored the effects of the large amounts of uncertainty that exists around the properties of self-clearance. No earlier models included self-clearance at all ([10], [16]). This neglect could result in biased estimates of vaccine impact, and potentially lead to suboptimal vaccine candidates being prioritised.

The aim of this work is to estimate the degree and direction of bias that may be resulting from the neglect of self-clearance in current models of tuberculosis vaccination.

## 2. Methods

### 2.1 Settings

Our analysis focuses on China and India. Conducting our modelling work on these countries allows us to evaluate how self-clearance may affect the potential impact of new vaccines in two countries that are key for the elimination of tuberculosis, and with different demographic and epidemiological characteristics. China and India are both high TB burden countries, accounting for about a third of the total global tuberculosis deaths in 2021 [1]. The epidemiology of tuberculosis and the demography differ in these two settings. India has a younger population, with a birth rate almost three times higher than China. Furthermore, China has experienced a more substantial reduction in tuberculosis incidence over the past few decades [17], with current estimates of incidence being 3–5 fold lower in China than in India.

### 2.2 Model and self-clearance structure

To estimate how self-clearance might affect the impact of novel tuberculosis vaccines, we adapted a compartmental age stratified dynamic Mtb transmission model that did include self clearance [8]. Ten compartments constituted the tuberculosis natural history, allowing for Mtb infection along a spectrum from uninfected to active clinical disease. Of these, five compartments referred to individuals with previous exposure to Mtb but without active disease:

- Infection-Fast (*I*_*F*_): individuals with a recent infection with Mtb (less than two years).
- Infection-Slow-Early 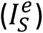 individuals last infected with Mtb in between two and nine years before, who have not cleared their infection.
- Infection-Slow-Late 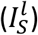: individuals last infected with Mtb more than nine years before, who have not cleared their infection.
- Uninfected-Cleared-Early 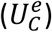 individuals last infected with Mtb in between two and nine years before, who have self-cleared their infection.
- Uninfected-Cleared-Late 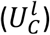 individuals last infected with Mtb more than nine years before,who have self-cleared their infection.

Figure 1 illustrates the flows into and from the Uninfected-Cleared compartments. The main difference between our model and the one in [8] is that we split the Uninfected-Cleared and Infection-Slow compartments into early (mean duration 7 years, and following a mean duration of 2 years in the Infection-Fast compartment) and late (starting an average of 9 years following last infection). This allowed us to calibrate the model to empirical data on the degree of protection against reinfection, which came from cohort studies where the follow-up period ended an estimated mean of nine years following infection [20]. This was done by varying the degree of protection against reinfection in people infected with *Mtb*, and assuming different levels of protection in people who had self-cleared (see Section 2.4.2).

**Figure 1.**
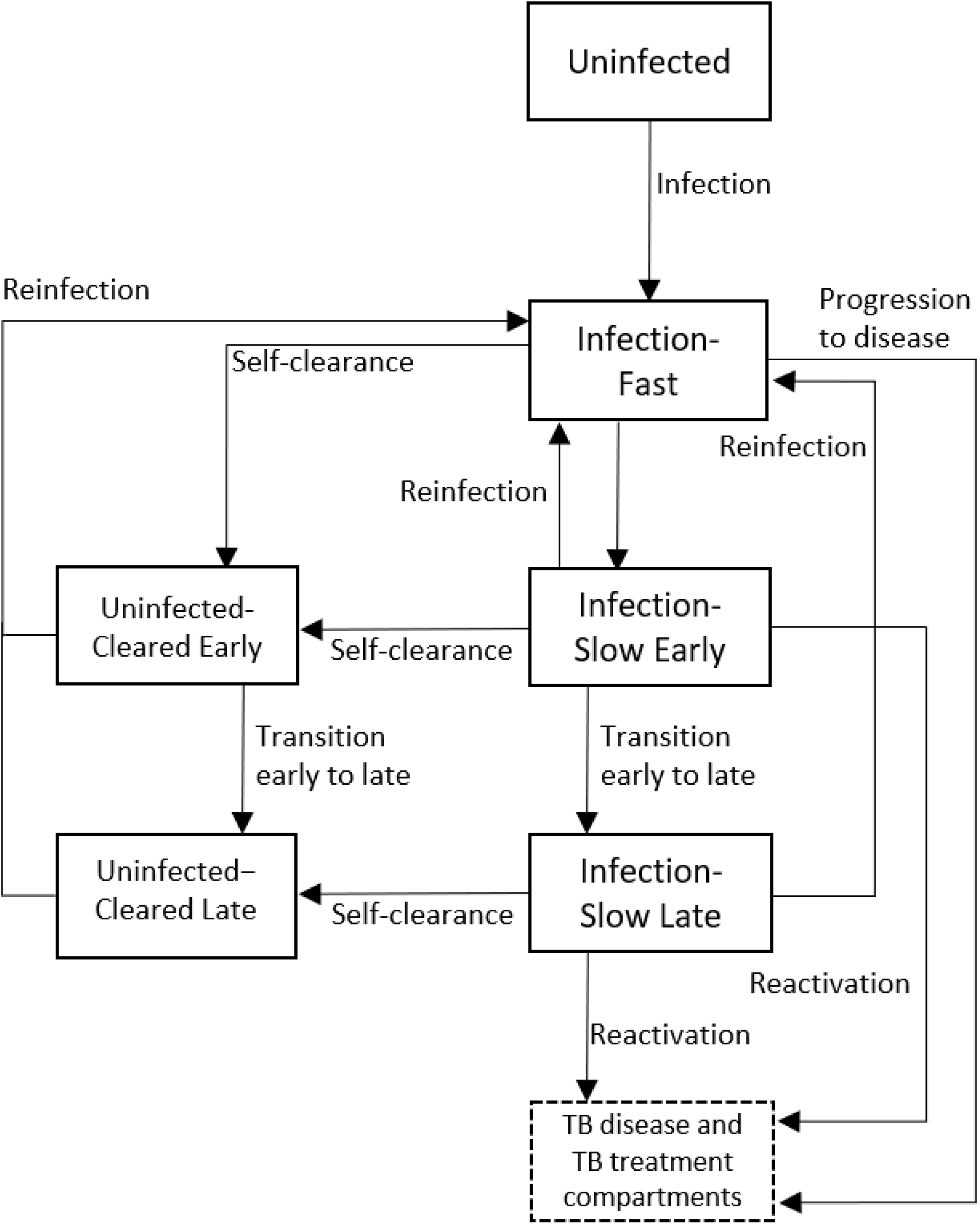
Diagram of the model structure showing parts of TB natural history related to infection and self-clearance in detail. *ϕ*_*S*_: rate of self-clearance from Infection-Slow Early to Uninfected-Cleared Late. The full model structure showing the disease and treatment compartments, is shown in Supplementary Material, Section A.

The properties of self-clearance were specified by the following two parameters in the natural history: the rate Φ_*S*_ at which individuals in Infection-Slow compartments self-cleared their infection, and the level of natural protection against reinfection that self-cleared individuals had, assumed to be *p*_*C*_ (0 ≤ *p*_*C*_ ≤ 1) times the natural protection against reinfection for individuals in the Infection-Slow compartments 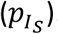

For details on the full model structure, please see Supplementary Material, Section A.

### 2.3 Vaccines

It is uncertain whether new TB vaccines will only work in uninfected people (no current infection vaccines, NCI), in people already infected with Mtb (current infection vaccines, CI), or in both (any infection vaccines, AI). Similarly, it is uncertain whether new TB vaccines will act to reduce the risk of infection (prevention of infection, POI), disease progression (prevention of disease, POD), or both (prevention of infection and disease, POID). We explored the impact of six different potential vaccines, consisting of the plausible combinations of the two characteristics (Table 1). A priori, we would expect that the inclusion of self-clearance in models will have limited effect on the impact of AI vaccines, as these vaccines have the same impact in all individuals, regardless of their infection status. We therefore focus mainly on NCI and CI vaccines in the results of this paper, and only consider the most extreme self-clearance scenarios for any infection vaccines (see Section 3.6).

**Table 1.** Characteristics of the six vaccines implemented.

| Abbreviation | Host infection status at time of vaccination required for efficacy | Effect type | Vaccine efficacy | Duration of protection |
| --- | --- | --- | --- | --- |
| NCI POI | No current infection (NCI) | Prevention of infection (POI) | 50% | Lifelong |
| NCI POID | No current infection (NCI) | Prevention of infection and disease (POID) | 50% | Lifelong |
| CI POD | Current infection (CI) | Prevention of disease (POD) | 50% | Lifelong |
| CI POID | Current infection (CI) | Prevention of infection and disease (POID) | 50% | Lifelong |
| AI POD | Any current infection (AI) | Prevention of disease (POD) | 50% | Lifelong |
| AI POID | Any current infection (AI) | Prevention of infection and disease (POID) | 50% | Lifelong |

Each vaccine was assumed to have a maximum efficacy of 50%, and was implemented with a one-time mass campaign for people aged 10 or older in 2025, and routine vaccination of 10-year-olds from 2025 to 2050. Both the mass campaign and the routine vaccinations were assumed to have a 70% coverage. Estimated vaccine impact was calculated as the tuberculosis incidence rate reduction in 2050 compared to a scenario where no new vaccine was implemented.

For more details on the implementation of the vaccine see Supplementary Material, Section C.

### 2.4 Self-clearance scenarios

In our central self-clearance scenario, we assumed a self-clearance rate equal to the median value provided in [15] (*ϕ*_*S*_= 0.033/year), a level of natural protection against reinfection in Uninfected-Cleared compartments equal to half the protection in the Infection-Slow compartments (*p*_*C*_ = 0.5), and a vaccine efficacy in self-cleared individuals equal to 25%, i.e. half the maximum efficacy assumed. To account for the uncertainty that exists in these values, we created a number of scenarios, varying them individually and in combination.

#### 2.4.1 Effect of different self-clearance rates on vaccine impact

To estimate the effect that different self-clearance rates may have on vaccine impact, we compared the scenario with no self-clearance with three scenarios where the self-clearance rate was set to the 2.5th percentile, the median and the 97.5th percentile provided in [15]. The other two characteristics, i.e. level of natural protection and vaccine efficacy in self-cleared individuals, were set to their mid values.

#### 2.4.2 Effect of different levels of natural protection in self-cleared individuals on vaccine impact

To estimate the effect that different levels of natural protection against reinfection in self-cleared individuals may have on vaccine impact, we compared the scenario with no self-clearance with three scenarios where the level of natural protection against reinfection in the Uninfected - Cleared compartments was set to 0%, 50% or 100% of the protection against reinfection in the Infection-Slow compartments. The other two characteristics, i.e. self-clearance rate and vaccine efficacy in self-cleared individuals, were set to their mid values.

#### 2.4.3 Effect of different levels of vaccine efficacy in self-cleared individuals on vaccine impact

To estimate the effect that different levels of vaccine efficacy in self-cleared individuals may have on vaccine impact, we compared the scenario with no self-clearance with three scenarios where the vaccine efficacy in self-cleared individuals was set to 0%, 25% and 50%. The other two characteristics, i.e. self-clearance rate and level of natural protection in self-cleared individuals, were set to their mid values.

#### 2.4.4 Maximum potential effect of including self-clearance in models of TB vaccination

To estimate the maximum effect that the inclusion of self-clearance in models may have on vaccine impact, we compared the scenario with no self-clearance with eight scenarios, corresponding to all possible combinations of the extreme values (minimum and maximum) of the three self-clearance characteristics (self-clearance rate, level of natural protection and vaccine efficacy in self-cleared individuals).

Furthermore, since the work in [15] was intended to provide a robust lower-bound of the self-clearance rate, we also estimated the maximum effect that self-clearance may have on vaccine impact if self-clearance rates higher than in [15] are considered. As the model was calibrated to the proportion of infected individuals, there is an upper bound on the proportion of all people who can be in the Uninfected-Cleared compartment. We were therefore able to generate a crude estimate of the maximum impact by extrapolating from the results of the low, medium, and high self-clearance rate scenarios. See Section D in Supplementary Material for full details.

### 2.5 Calibration process

For each country and each self-clearance scenario (with the exception of scenarios varying the vaccine efficacy in self-cleared people, where the baseline fit does not change), we re-calibrated the model to same epidemiologic data using history matching with emulation using the hmer R package ([18], [19]), generating at least 1000 fitted parameter sets. Results are presented as the median and 95% plausible range, calculated as the 2.5 and 97.5 percentile across the 1000+ available parameter sets.

The model for each country and scenario was calibrated to 9-12 calibration targets in 2019, including the tuberculosis case notification rate (overall and by age), tuberculosis mortality rate (overall), and the proportion of prevalent tuberculosis that is subclinical. We also calibrated the model to data on the prevalence of latent Mtb infection in individuals ages 15-99 (in 2000 for China and 2021 in India). To ensure that the evolution of the tuberculosis epidemic was captured, we matched to time trends in the tuberculosis disease prevalence in China (1990 and 2010), and to time trends in tuberculosis incidence in India (2000 and 2019). Finally, we matched to the estimate of the degree of protection against reinfection in latent individuals provided in [20]. Full details on the calibration targets are given in Section B in the Supplementary Material.

## 3 Results

### 3.1 Baseline scenarios

We successfully calibrated each self-clearance scenario for the two countries of interest, generating at least 1000 fits per scenario. Figure 2 shows how different values of the self-clearance rate and of the natural protection against reinfection in self-cleared individuals, affected the proportion of individuals in the various compartments, and the distribution of the flows into the Infection-Fast compartment in China in 2025. The corresponding plots for India are very similar and can be found in Supplementary Material, Section B. Since all individuals progressing to active disease need first to have passed through the Infection-Fast compartment, analysing how the flows into the Infection-Fast compartment vary across self-clearance scenarios will help us understand how incidence is affected by different parameterisations of self-clearance.

**Figure 2.**
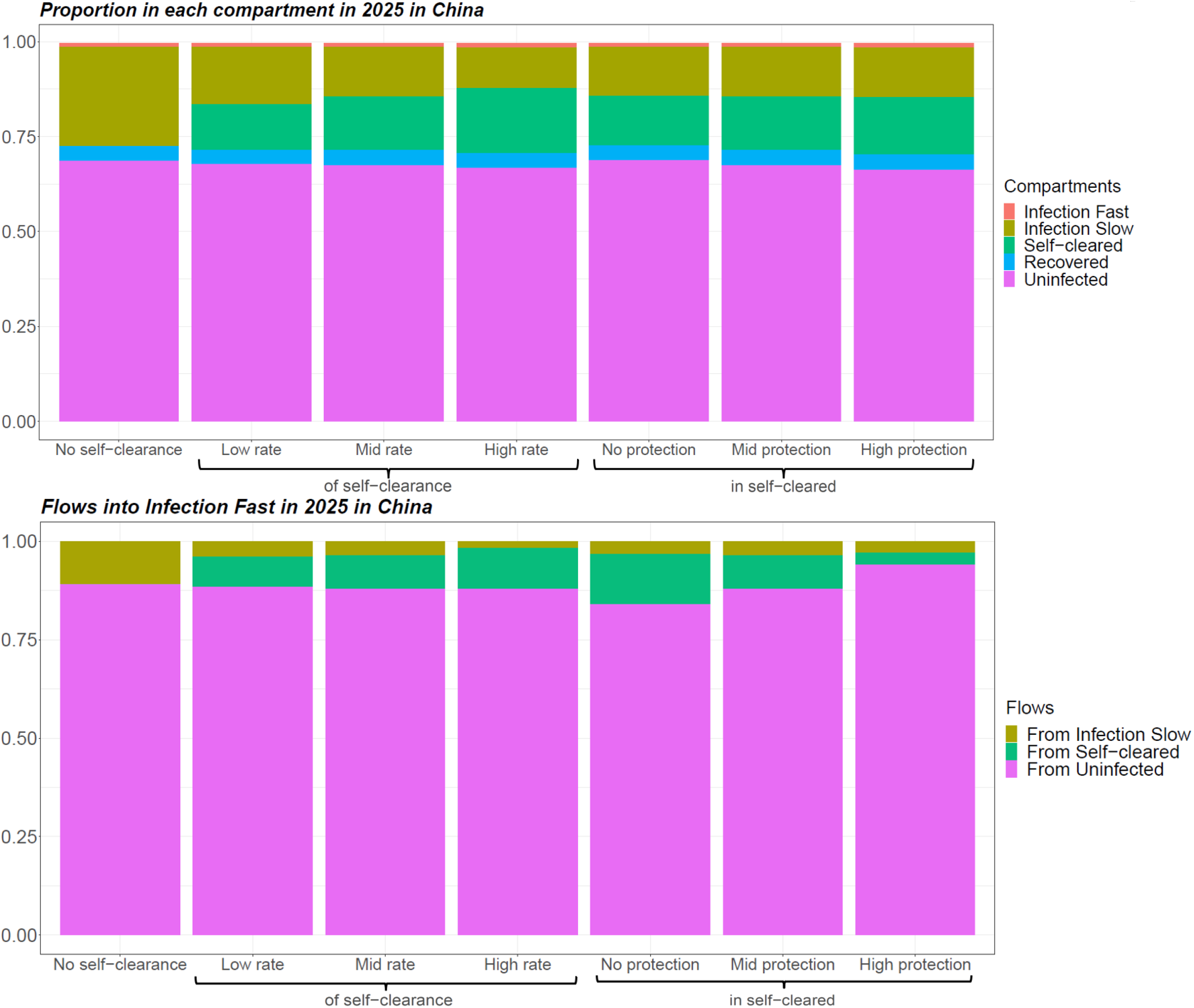
Proportion of individuals in each natural history state and distribution of flows into Infection-Fast in 2025 for different baseline scenarios in China, averaged across model runs. In scenarios with self-clearance, when the rate of self-clearance or the level of protection in Uninfected-Cleared individuals are not indicated, they are fixed at their mid value.

As expected, we see that higher self-clearance rates result in higher proportions of self-cleared individuals. On the other hand, varying the level of natural protection against reinfection in self-cleared individuals does not substantially change the proportion of individuals in the various compartments. There are three types of flows into the Infection-Fast compartment: from the Infection-Slow compartments, from the Uninfected-Cleared compartments and from the Uninfected-Naive compartment. In both countries, the size of the flow from Uninfected-Cleared compartments is slightly higher when the self-clearance rate is higher and substantially lower when the natural protection against reinfection in Uninfected-Cleared individuals is lower. Furthermore, the proportion of all flows into the Infection-Fast compartment that come from the Uninfected-Cleared compartments is always below 15% in both countries: this suggests that people who are self-cleared cannot directly affect more than 15% of the tuberculosis incidence.

### 3.2 Effect of self-clearance on vaccine impact (central scenario)

In the no-self-clearance scenario and central scenarios respectively, the no current infection, prevention of infection vaccine resulted in an relative incidence reduction in 2050 of 35.3% and 38.5% in China and 29.1% and 30.1% in India; the no current infection, prevention of infection and disease vaccine in a reduction of 46.5% and 50.7% in China and 39.7% and 41% in India; the current infection, prevention of disease vaccine in a reduction of 27.9% and 27.5% in China and 25.2% and 25.8% in India; and the current infection, prevention of infection and disease vaccine in a reduction of 29.2% and 29.2% in China and 26.2% and 26.4% in India (Figure 3). Overall in both countries the largest relative incidence reduction was observed for no current infection vaccines: this is in accordance with the fact that the flow into Infection Fast coming from uninfected individuals (on whom no current infection vaccines are effective) was substantially larger than the flow coming from infected individuals (on whom current infection vaccines are effective) (Figure 2).

**Figure 3.**
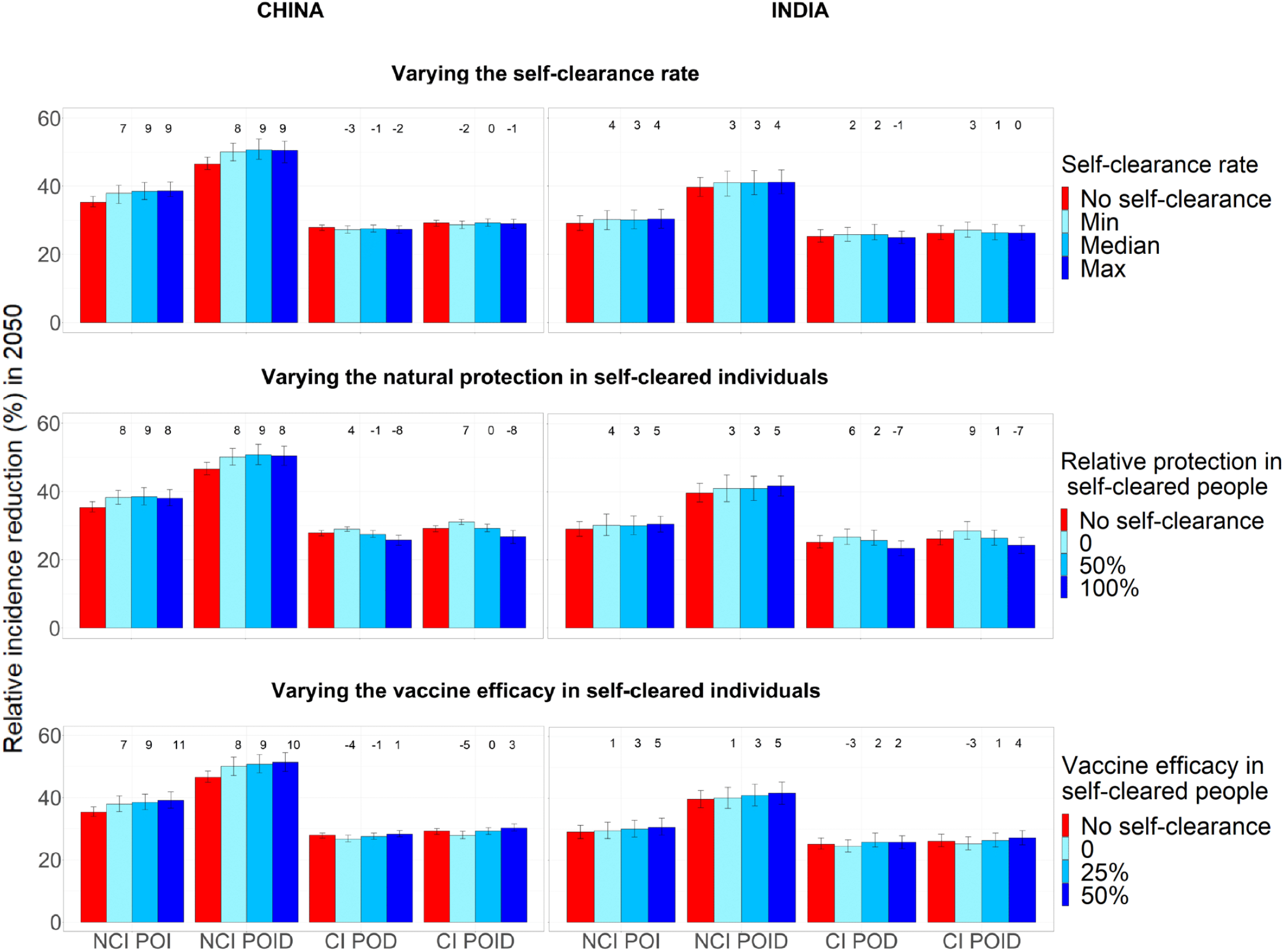
Relative incidence reduction (%) in 2050 compared to a scenario with no vaccination, when each of the three self-clearance characteristics is varied separately for no current infection vaccines and current infection vaccines (China on the left, India on the right). For each plot, the two unshown characteristics are fixed to their mid value. The height of the columns show the median percentage reduction in incidence in 2050 compared to the scenario where no vaccine is implemented, with vertical bars indicating the 95% confidence interval. The numbers on top of each column correspond to the relative change in incidence reduction compared to the no self-clearance scenario. NCI: no current infection, CI: current infection, POI: prevention of infection, POD: prevention of disease, POID: prevention of infection and disease.

### 3.3 Effect on vaccine impact of different self-clearance rates

Figure 3 shows how estimated relative reduction in tuberculosis incidence 2050, compared to the baseline no vaccination scenario, varies when varying the three self-clearance characteristics (self-clearance rate, level of natural protection and vaccine efficacy in self-cleared individuals) separately. With the exception of the different vaccine efficacy scenarios, the model was calibrated separately to each scenario. Minor differences between scenarios in estimated vaccine impact may therefore reflect minor, chance differences between the baseline fitted scenarios, and not genuine differences in vaccine impact, particularly where trends are not observed.

Varying the self-clearance rate has only a small effect on estimated vaccine impact, for any vaccine in either country. The maximum relative increase in vaccine impact compared to the no self-clearance scenario when only varying the self-clearance rate, was 9% in China and 4% in India, while the maximum relative decrease was 3% in China and 1% in India. This is in accordance with Figure 2: since the distribution of the flows into Infection-Fast changes little with the value of the self-clearance rate, we would not expect the impact of vaccines on tuberculosis incidence to be strongly affected by the rate of self-clearance.

### 3.4 Effect of different levels of natural protection in self-cleared individuals, on vaccine impact

The maximum relative increase in vaccine impact when varying the level of natural protection against reinfection in self-cleared individuals, was 9% in both countries, while the maximum relative decrease was 8% in China and 7% in India. Increasing the level of natural protection against reinfection in self-cleared individuals decreased the impact of current infection vaccines. This is because as the level of protection was increased, the proportion of the flow into Infection-Fast that comes from the Self-Cleared compartments decreased (from 12.7% to 2.8% in China, and from 12.9% to 4.2% in India), and the proportion that comes from Uninfected-Naive increased (from 84.3% to 94.3% in China, and from 80.4% to 91.6% in India) (Figure 2). As we assumed that current infection vaccines offered partial protection to self-cleared individuals, and none to uninfected individuals, this resulted in a lower estimated vaccine impact in scenarios with a higher level of natural protection in self-cleared individuals.

The results do not show the opposite trend for no current infection vaccines: an increase in vaccine impact as we increase the level of natural protection. The reason for this is that the relative change in the percentage of the flow into Infection-Fast coming from individuals in which the vaccines are effective is small (from 90.6% to 95.6% in China, and from 88.4% to 93.7%), and the effect on vaccine impact is therefore masked by minor differences between the calibrated baseline scenarios.

### 3.5 Effect of different levels of vaccine efficacy in self-cleared individuals on vaccine impact

The maximum relative increase in vaccine impact when varying the level of vaccine efficacy in self-cleared individuals was 11% China and 5% in India, while the maximum relative decrease was 5% in China and 3% in India. As expected, increasing the vaccine efficacy in self-cleared individuals, increases the estimated vaccine impact for all vaccine types. The increase in estimated impact for no current infection vaccines was substantially higher in China, compared to India. The main reason for this is to be found in the demographic differences between the two countries. Since only a very small percentage of 10 year-olds are in the self-cleared state (less than 1% both in China and India), self-clearance has a larger effect on the mass vaccination campaign than the routine vaccination of children. The effects of the mass vaccination are diluted much more quickly in India than China, due to the much higher birth rate, explaining the difference observed in Figure 3 third row.

### 3.6 Maximum potential effect of including self-clearance in models of TB vaccination

Figure 4 shows the maximum relative decrease and increase in tuberculosis incidence reduction in 2050 when we varied all three self-clearance characteristics at once. The maximum relative increase for no current infection vaccines in our model was 12% in China and 8% in India. For current infection vaccines, the maximum relative increase was 12% in China and 15% in India, while the maximum relative decrease was 14% in China and 11% in India. Unsurprisingly, including self-clearance in the model only increased the impact of no current infection vaccines, as it increased the number of people in which vaccines work from people who have never been infected at the time of vaccination only, to also include people who have self-cleared (Figure 2, first row). By the same logic, we may suppose that including self-clearance could only decrease the impact of current infection vaccines, however that is not what we observe: modelling self-clearance increases estimated vaccine impact in a number of scenarios, with the greatest increases found where self-cleared people have no protection against reinfection. The main reason is that a higher proportion of the flow into the Infection-Fast compartment comes from compartments where current infection vaccines can offer some protection (i.e. the Infection-Slow and self-cleared compartments) in scenarios where we assume that self-cleared people have reduced or no protection against reinfection, compared to scenarios with no self-clearance (Figure 2, second row). Additionally, 26-35% of the vaccine doses are delivered through routine vaccination of 10-year-olds in the model, a group in which the prevalence of latent infection is 4-10 fold higher than the prevalence of self-cleared infection, even in the highest self-clearance rate scenarios.

**Figure 4.**
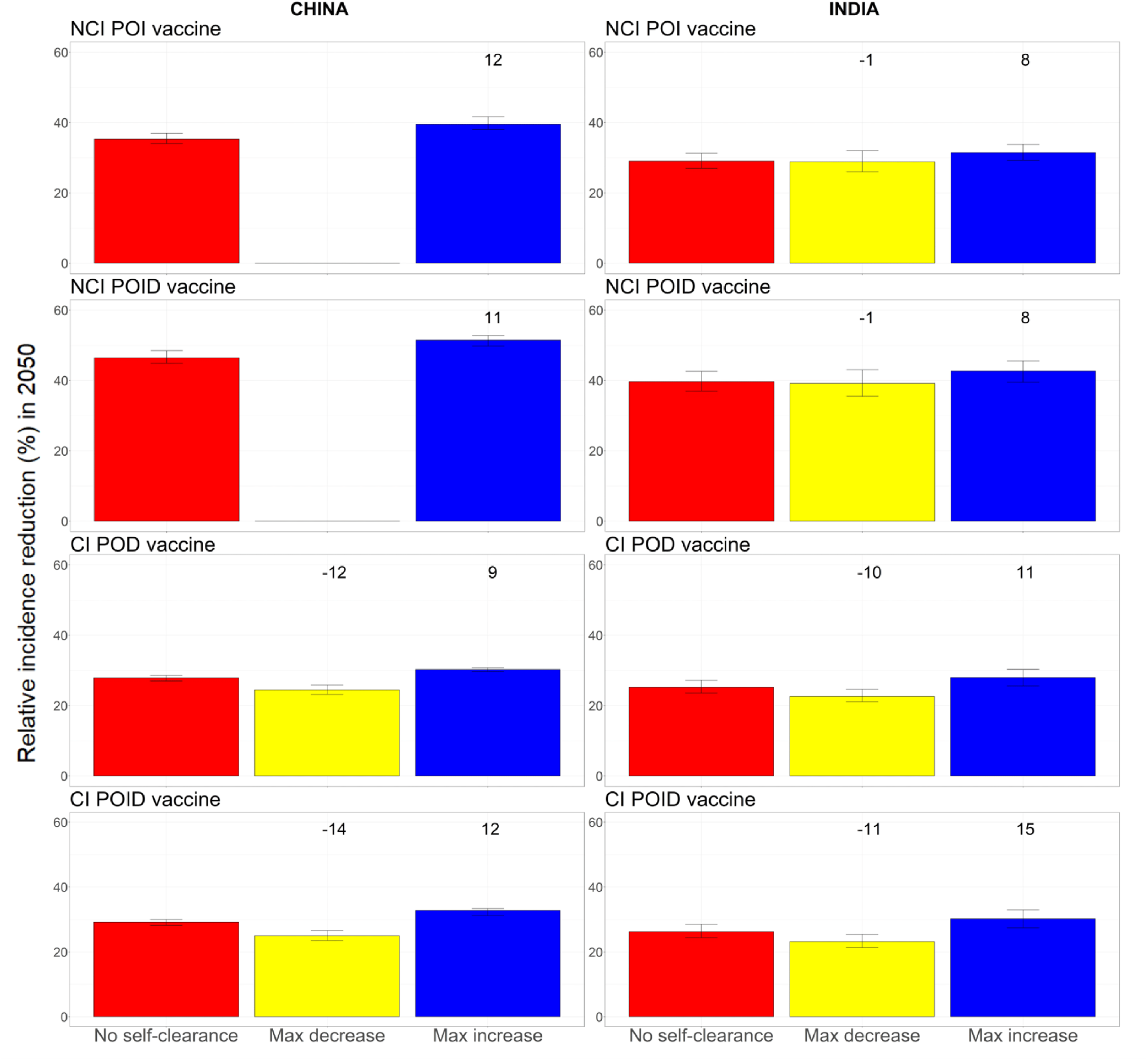
Relative incidence reduction (%) in 2050 compared to a scenario with no vaccination, when all three self-clearance characteristics are varied at once for no current infection vaccines and current infection vaccines (China on the left, India on the right). The height of the columns show the median percentage reduction in incidence in 2050 compared to the scenario where no vaccine is implemented, with vertical bars indicating the 95% confidence interval. The numbers on top of each column correspond to the relative change in incidence reduction compared to the no self-clearance scenario. For no current infection vaccines, the yellow columns are missing in China, as no explored scenario resulted in a decreased vaccine impact. NCI: no current infection, CI: current infection, POI: prevention of infection, POD: prevention of disease, POID: prevention of infection and disease.

See supplementary Material, Section D, for details on the specific scenarios in which each maximum decrease and increase were achieved for each vaccine.

As the empirical estimate of self-clearance rates used in the model was designed to be a plausible lower bound [15], we also estimated the maximum potential effect of self-clearance on vaccine impact for higher self-clearance rates. Even when assuming that self-clearance occurs instantly from the Infection states, the estimated maximum relative increase for the impact of no current infection vaccines was never higher than 16%, while the estimated maximum relative increase/decrease for the impact of current infection vaccines was never higher than 25% (cf. Table 2). For the complete analysis see Supplementary Material, Section D.

**Table 2.** Upper bounds on the maximum increase and decrease in tuberculosis incidence reduction in 2050, compared to the no self-clearance scenario, that can result when allowing self-clearance rates higher than those provided in [15]. These values are obtained in the extreme case where there are no individuals in Infection-Slow compartments, so they should be regarded as strong, limit values, which are not realistic. NCI: no current infection, CI: current infection, POI: prevention of infection, POD: prevention of disease, POID: prevention of infection and disease.

|  | China |  | India |  |
| --- | --- | --- | --- | --- |
|  | Max increase | Max decrease | Max increase | Max decrease |
| NCI POI | 12% | 0% | 16% | 1% |
| NCI POID | 11% | 0% | 16% | 1% |
| CI POD | 14% | 21% | 17% | 19% |
| CI POID | 20% | 25% | 25% | 22% |

### 3.7 Effect of self-clearance on the impact of any infection vaccines

As hypothesised, the inclusion of self-clearance has little effect on the impact of any infection vaccines, with the tuberculosis incidence reduction in 2050 being increased by a maximum of 6% in China and 3% in India, relative to the scenario with no self-clearance (see Supporting Material, Section E for full results). These small changes are likely to result from chance differences between the baseline fitted scenarios, and not genuine differences in vaccine impact.

## 4. Discussion

Our results suggest that the neglect of self-clearance in mathematical models of tuberculosis is not leading to substantial bias in estimates of vaccine impact, with maximum relative increases or decreases in impact of up to 15% only. As expected, modelling self-clearance had little effect on the estimated impact of any infection vaccines, as these vaccines are assumed to work in all people, regardless of infection status. Not including self-clearance in models may mean that we are slightly underestimating the impact of no current infection vaccines, slightly under-or over-estimating the impact of current infection vaccines, and slightly underestimating the increased impact of no current infection vaccines compared to current infection vaccines. Underestimation of the impact of no current infection vaccines is likely to be higher for vaccine roll-out scenarios that target older age groups, compared to children or adolescents only, and for countries with lower birth rates.

This paper presents some of the first work exploring how the inclusion of self-clearance in mathematical models affects estimates of intervention impact. We demonstrate that self-clearance does not substantially affect estimates of vaccine impact, however it is still to be investigated how the neglect of self-clearance in TB models may affect estimates of impact for other interventions, (e.g. preventative treatment or different screening strategies). Based on our analysis, a relatively small proportion of incident disease occurs in (previously) self-cleared individuals, in two high burden countries with very different demographic and epidemiological characteristics. Taking this into consideration, it seems unlikely that neglecting self-clearance in tuberculosis models would lead to substantially biased estimates of the impact of interventions targeting the overall population. The impact of self-clearance might, however, be higher for interventions that target or only work in subgroups with a substantially larger proportion of self-cleared individuals than in the entire population.

It should be noted that the aim of this work was to explore the effect of including self-clearance in models of vaccine impact, not to obtain estimates of vaccine impact in differing vaccine and implementation scenarios. In all implemented scenarios, we assumed that vaccines would confer lifelong protection (against infection and/or progression to disease) and that they were delivered with a mass campaign for people aged 10 or more, followed by the routine vaccination of 10-year-olds. These assumptions were made in order to maximise the potential vaccine impact, and the effect of including self-clearance on vaccine impact. Similarly, we did not include some details in the model that could have improved estimates of vaccine impact, but had little effect on how modelling self-clearance altered impact (e.g. differences in treatment mortality and completion probabilities between the public and private sector in India).

In this work we chose not to model any countries where HIV plays an important role for the epidemiology of TB. This is because of the even greater levels of uncertainty around characteristics of self-clearance of Mtb infection in people living with HIV. However, it seems plausible to assume that being HIV positive would reduce people’s ability to self-clear their Mtb infection. This suggests that neglecting self-clearance in models for countries with a significant co-interaction of tuberculosis and HIV may also not result in substantially biased estimates of vaccine impact.

There are a number of limitations to our work. First, in our main analysis we modelled self-clearance rates according to the estimates in [15], which were meant to provide a robust lower bound for the self-clearance rate. To account for the fact that self-clearance rates may be higher than these estimates, we also explored higher self-clearance rates through statistical estimation of their potential values. We showed that the estimated maximum relative increase in vaccine impact compared to the no self-clearance scenario for no current infection vaccines was never higher than 16%, and the estimated maximum relative increase/decrease for current infection vaccines was never higher than 25%. These values strengthen the message that neglecting self-clearance does not lead to substantially biased estimates of vaccine impact. Another limitation is that modelling different baseline scenarios inevitably leads to chance differences between scenarios. These differences are minor only, and are unlikely to have had any real impact on our conclusions, however they may have masked minor differences between scenarios.

TB natural history, and how best to represent it in mathematical models, is an area of active research. Future work may indicate that other changes to model structure could improve our representation of TB. For instance, modifications may be necessary if empirical data suggest that vaccine efficacy varies between people who have never been exposed to *Mtb*, and people who have been exposed without developing a detectable immune response. Changes in how these aspects of natural history are simulated could lead to higher or lower estimates of vaccine efficacy.

In conclusion, our work suggests that the neglect of self-clearance in mathematical models of TB vaccines does not result in substantially biased estimates of TB vaccine impact. It may, however, mean that we are slightly underestimating the relative advantages of vaccines that work in uninfected individuals only compared to those that work in infected individuals.

## Data Availability

All data produced are available online (WHO database and UN population estimates).

