## Supplementary Material for "Is neglect of self-clearance biassing TB vaccine impact estimates?"

### A. Model details

Section A describes details regarding the epidemiological model based off of Clark et al. [8]. A compartmental deterministic dynamic model of *Mycobacterium tuberculosis* (*Mtb*) transmission and progression was developed, structured by the following core dimensions: 82 age compartments, 10 tuberculosis natural history compartments, two socio-economic status (SES) compartments defined by access-to-care.

#### Age dimension

Age was modelled in single years from ages 0 to 79 and aggregated into two categories for ages 80 to 89, and ages 90 to 99. Births and ageing occurred at the beginning of each year.

#### Natural history dimension

The tuberculosis natural history was represented using ten different compartments, as shown in Figure A. Natural history compartments and parameters are described in Tables A and B.

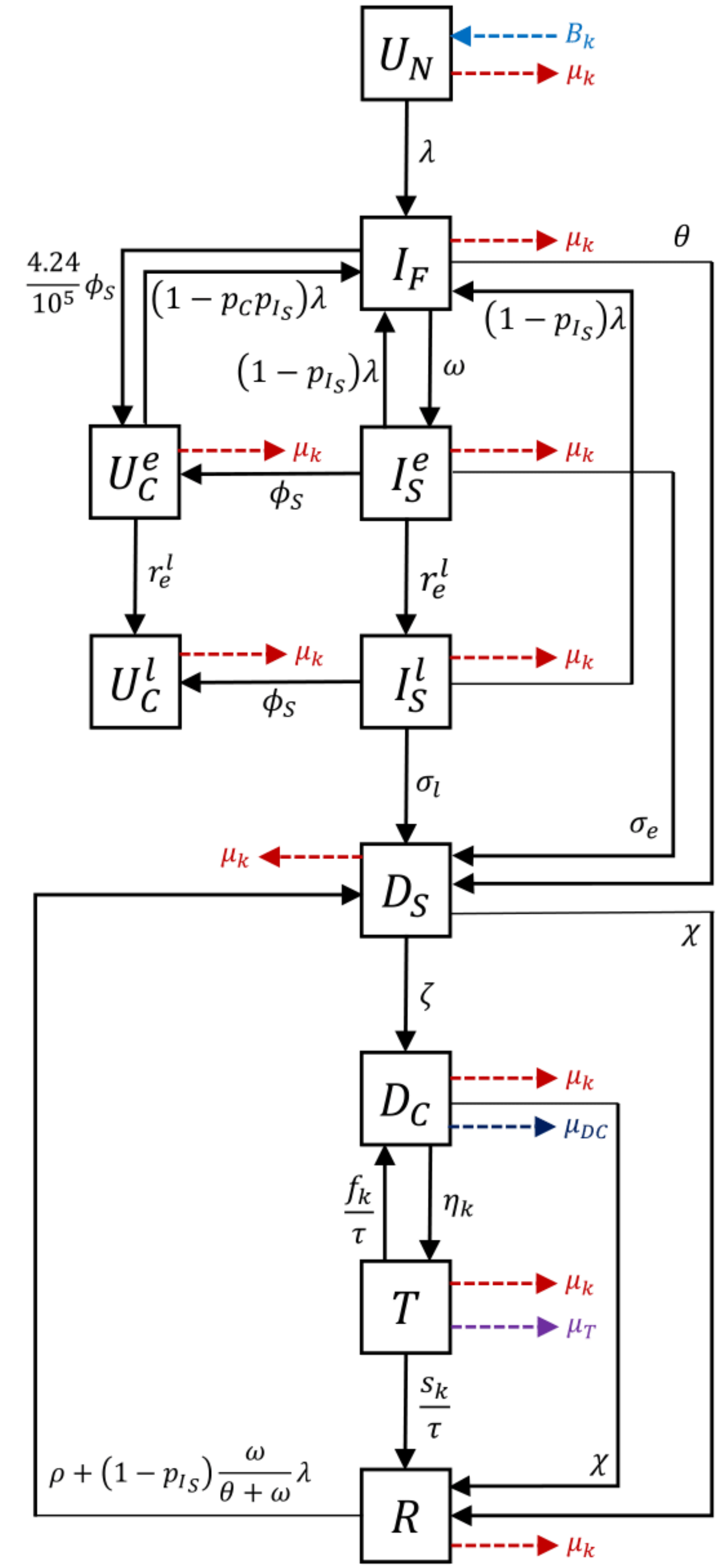


Figure A: Tuberculosis natural history in our model

Table A. Description of natural history compartments

| Symbol | Compartment Name | Compartment Description |
|---|---|---|
| $U_N$ | **Uninfected - Naive** | Individuals with no previous *Mtb* infection |
| $\boldsymbol{U_C^e}$ | **Uninfected - Cleared Early** | Individuals *with* previous exposure/infection who have self-cleared. Early indicates that the infection occurred at most 9 years before. No viable bacteria remaining. |
| $\boldsymbol{U_C^l}$ | **Uninfected - Cleared Late** | Individuals *with* previous exposure/infection who have self-cleared. Late indicates that the infection occurred at least 9 years before. No viable bacteria remaining. |
| $\boldsymbol{I_F}$ | **Infection - Fast** | Infected individuals who were last infected at most 2 years before |
| $\boldsymbol{I_S^e}$ | **Infection - Slow Early** | Individuals last infected with Mtb in between two and nine years before, who have not cleared their infection. |
| $\boldsymbol{I_S^l}$ | **Infection - Slow Late** | Individuals last infected with Mtb more than nine years before, who have not cleared their infection. |
| $D_S$ | **Subclinical Disease** | Individuals with active, infectious TB disease, who do not report any of the four WHO TB symptom-screen symptoms |
| $D_C$ | **Clinical Disease** | Individuals with active, infectious TB disease, who report any of the four WHO TB symptom-screen symptoms |
| $T$ | **On-treatment** | Individuals detected from Disease Clinical and started on treatment |
| $R$ | **Resolved** | Completed treatment or naturally cured from Disease Clinical and Disease Subclinical |

Table B. Description of the parameters used in the natural history dimension.

| Parameter Description | Units | Symbol |
|---|---|---|
| ***Births*** | | |
| Births | Overall, per year | $B_k$ |
| ***Natural history*** | | |
| Rate of natural cure from Clinical Disease ($D_C$) and Subclinical Disease ($D_S$) | Per person, per year | $\chi$ |
| Rate of progression to disease from Infection-Fast | Per person, per year | $\theta$ |
| Age scaling parameter for the rate of fast progression ($\theta$) | - | $j_1$ |
| Rate of reactivation from Infection – Slow Early | Per person, per year | $\sigma^e$ |
| Rate of reactivation from Infection – Slow Late | Per person, per year | $\sigma^l$ |
| Age scaling parameter for rate of reactivation from Infection – Slow ($\sigma$) | - | $j_2$ |
| Rate of relapse from Resolved (R) | Per person, per year | $\rho$ |
| Age scaling parameter for rate of relapse from Resolved ($\rho$) | - | $j_3$ |
| Rate of progression from Subclinical Disease to Clinical Disease | Per person, per year | $\zeta$ |
| Rate of self-clearance from Infection – Slow compartments to Uninfected – Cleared compartments | Per person, per year | $\phi_S$ |
| Rate of self-clearance from Infection – Fast compartments to Uninfected – Cleared compartments | Per person, per year | $0.0000424\phi_S$ |
| Probability of transmission per infectious contact | - | $p_T$ |

| | | |
|---|---|---|
| Force of infection | Per year | $\lambda$ |
| Rate from Infection - Fast to Infection- Slow Early | Per person, per year | $\omega$ |
| Multiplier for TB prevalence | - | - |
| Rate of transition from Infection - Slow Early to Infection - Slow Late and from Uninfected - Cleared Early to Uninfected - Cleared Late. | Per person, per year | $\boldsymbol{r_e^l}$ |
| Relative protection against reinfection in Uninfected - Cleared compartments, compared to $p_{I_S}$ | - | $\boldsymbol{p_C}$ |
| ***Mortality*** | | |
| Mortality rate from untreated clinical TB | Per person, per year | $\mu_{DC}$ |
| Background mortality rate | Per person, per year | $\mu_k$ |
| TB Mortality rate on treatment | Per person, per year | $\mu_T$ |
| Age scaling parameter for mortality rates | - | $s_{Age}$ |
| ***Treatment*** | | |
| Rate of treatment initiation | Per person, per year | $\eta$ |
| Treatment duration | Year | $\tau$ |
| Age scaling parameter for rate of treatment initiation ($\eta$) | - | $j_4$ |
| Rate of treatment completion | Per person, per year | $\frac{s}{\tau}$ |
| Rate of treatment non-completion | Per person, per year | $\frac{f}{\tau}$ |
| ***Protection*** | | |
| Access-to-care parameter | - | $p_E$ |
| Protection from reinfection offered by Infection – Slow and Resolved compartments | - | $p_{I_S}$ |

### Force of infection

The equation for the age-specific force of infection ($\lambda_j$) is given below. Clinically, tuberculosis can present as pulmonary tuberculosis which impacts the lungs, and/or extrapulmonary tuberculosis (EPTB) which occurs in sites other than the lungs, and is predominantly non-infectious. The WHO tuberculosis estimates which we calibrated the model to include both EPTB and pulmonary tuberculosis. We therefore reduced the simulated force of infection by the estimated proportion of incident cases that are EPTB to account for the fact that they are not infectious.. We also reduced the simulated force of infection to account for the reduced infectiousness of subclinical disease compared to clinical disease.

$$\lambda_j = p_T \times \sum_{y=1}^{n_{ygroups}} C[m,y] \times \left( \frac{(1-ep)(TD_{C_y} + rTD_{S_y})}{N_y} \right)$$

Where

$$\mathrm{N}_y = \sum_{j=j_{min}}^{j_{max}} U_{N_j} + U_{C_j}^e + U_{C_j}^l + I_{F_j} + I_{S_j}^e + I_{S_j}^l + D_{S_j} + D_{C_j} + T_j + R_j$$

$$TD_{C_y} = \sum_{j=j_{min}}^{j_{max}} D_{C_j} \qquad TD_{S_y} = \sum_{j=j_{min}}^{j_{max}} D_{S_j}$$

| Parameter | Definition |
| --- | --- |
| $j$ | Age of individual (in years) |
| $y$ | Age group of contact |
| $n_{ygroups}$ | Number of contact age groups |
| $p_T$ | Accounting for the probability of transmission per infectious contact |
| $m$ | Age group of individual |
| $C[m, y]$ | Contact rate between individual of age group $m$ and contact of age group $y$ |
| $ep$ | Average proportion of tuberculosis cases that are extrapulmonary |
| $r$ | Proportional reduction in infectiousness from subclinical disease |
| $TD_{C_y}$ | Total population in a clinical disease class in age group $y$ |
| $TD_{S_y}$ | Total population in a subclinical disease class in age group $y$ |
| $N_y$ | Total population alive in age group $y$ |
| $j_{min}$ | Minimum age $j$ within age group $y$ |
| $j_{max}$ | Maximum age $j$ within age group $y$ |

#### Access to care dimension

The access to care dimension contains 2 compartments: high-access-to-care, representing the top 3 quintiles (60% of the population in each country) and low-access-to-care, representing the bottom 2 quintiles (40% of the population in each country). We assumed that there was no transition between the high- and low-access-to-care compartments, as well as assuming random mixing between the high-access-to-care and low-access-to-care compartments.

To constrain relative burden between access-to-care compartments, we calibrated the relative tuberculosis prevalence in the high-access-to-care compartment to the low-access-to-care compartment in 2019. The calibration target, 0.674, was calculated as a weighted average from ten studies [21]–[30], with lower and upper bounds (0.575–0.801) representing the 25th and 75th percentiles of the datasets.

To incorporate access to care into our model, we assume that the differences in tuberculosis burden between compartments are due to differences in the force of infection, the rate of care-seeking (i.e., tuberculosis treatment initiation), and the rate of progression to tuberculosis following infection. We assume relative to the low-access-to-care strata, the high-access-to-care strata has a reduced force of infection per contact, an increased rate of treatment initiation, and a reduced rate of progression. Differential burden was implemented by introducing a new parameter $p_E$, such that $p_E \in [0,1]$, for the high-access-to-care and $p_E = 0$ for the low-access-to-care compartment. This new parameter was included within the model natural history structure as described in Table 3 and was fitted during calibration.

| | Access-to-Care |
|---|---|
| **Force of infection** | $p_T \times \sum_{y=1}^{n_{ygroups}} (1-p_E) \times C[m,y] \times \left(\frac{(1-ep)(TD_{C_y}+rTD_{S_y})}{N_y}\right)$ |
| **Treatment Initiation Rate** | $\frac{\eta_{j,k}}{(1-p_E)}$ |
| **Rate of Tuberculosis Progression** | $(1-p_E) \times \theta_j$<br>$(1-p_E) \times \sigma_j$<br>$(1-p_E) \times \rho_j$ |

Table C. Implementation of the access-to-care parameter $p_E$

### Tuberculosis treatment initiation

Tuberculosis treatment was implemented in the model from 1960, aligned roughly with the discovery and widespread use of rifampicin. The simulated rate of starting treatment increases following a sigmoid curve to 2019 (Figure B). The treatment initiation rate parameter, $\eta_j$, represents the age specific rate of treatment initiation from the clinical disease compartment. During calibration, we varied a country-specific value for $\eta_j$ which was sampled between 0 and 1. $\eta_j$ was then multiplied by an age scaling parameter for children, $j_4$, also sampled between 0 and 1, to ensure that the treatment initiation rate in children was less than in adults. This was then multiplied by the value of the sigmoid curve at each year. The model was calibrated to the country-specific notification rate in 2019 overall and by age reported by the WHO.

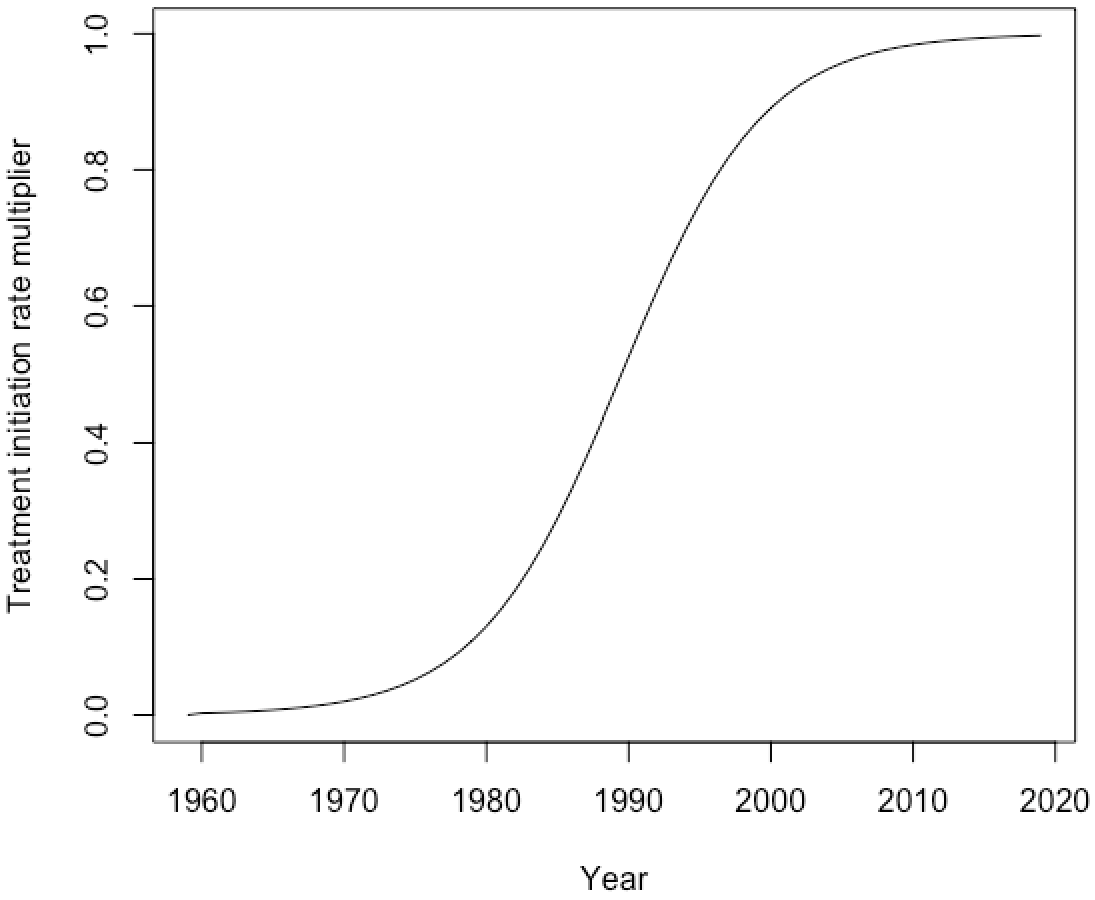


Figure B. Sigmoid curve representing the scale-up in tuberculosis treatment from 1960-2019

### Tuberculosis treatment outcome

In addition to background mortality, there are three possible exits from the on-treatment compartment: treatment completion, which progresses to the resolved compartment; treatment non-completion, which returns to the clinical disease compartment; and on treatment mortality, which counts toward tuberculosis mortality. To account for the variability in tuberculosis treatment outcomes and possible underreporting of on-treatment mortality, we used the following country-specific process:

1. For each country separately, the proportion of treatment completions was calculated and averaged over the years of available data from WHO.
2. A value for child treatment mortality $k_{A_0}$ was sampled between 0 and $2 \times Average\ Reported\ Treatment\ Mortality$. The average reported treatment mortality was multiplied by 2 to give an upper bound in the case of unreported data.
3. The age multiplier, $s_{age}$, was sampled from $(0,1)$, and multiplied by $k_{A_0}$ to calculate the adult treatment mortality $k_{A_{15}}$.
4. The success and failure rates per year were calculated as in Table D.
5. Each of the parameters in Table 1 were divided by the treatment duration $\tau$ to obtain the on-treatment mortality rate per year, on-treatment completion rate per year, and on-treatment non-completion rate per year.

| Parameter | Adults | Children |
|---|---|---|
| $\kappa_j$<br>On-treatment mortality fraction | $\kappa_{A0} \times S_{Age}$ | Sample $\kappa_{A0}$ from<br>0 to 2 x Average mortality on-treatment |
| $s_j$<br>On-treatment completion fraction | $(1-\kappa_{A15})\mathrm{SFR}$ | $(1-\kappa_{A0})\mathrm{SFR}$ |
| $f_j$<br>On-treatment non-completion fraction | $(1-\kappa_{A15})(1-\mathrm{SFR})$ | $(1-\kappa_{A0})(1-\mathrm{SFR})$ |

Table D. Calculating treatment outcome parameter values for adults and children.

### B. Calibration

#### Calibration targets

Table E shows the calibration targets with their mean, their 95% confidence intervals (in brackets) and the sources used to calculate them.

| | China | | | | India | | | |
|---|---|---|---|---|---|---|---|---|
| **Target Description** | **Year** | **Age group** | **Ranges** | **Sources** | **Year** | **Age group** | **Ranges** | **Sources** |
| Proportion of prevalent TB that is subclinical | 2019 | 0-99 | 0.504 [0.361-0.797] | [31] | 2019 | 0-99 | 0.504 [0.361-0.797] | [31] |
| TB prevalence ratio by access-to-care | 2019 | 0-99 | 0.674 [0.575-0.801] | [21]–[30] | 2019 | 0-99 | 0.674 [0.575-0.801] | [21]–[30] |
| TB disease prevalence (per 100,000 population) | 1990 | 15-99 | 635 [603-666] | [17] | 2021 | 15-99 | 427.5 [372-483] | [32] |
| TB disease prevalence (per 100,000 population) | 2010 | 15-99 | 459 [433-484] | [17] | - | - | - | - |
| TB mortality (per 100,000 population/year) | 2019 | 0-99 | 2.3 [2.1-2.6] | [33] | 2019 | 0-99 | 33 [30-35] | [33] |
| TB notifications (per 100,000 population/year) | 2019 | 0-14 | 2.6 [2.1-3.1] | [34] | 2019 | 0-14 | 40 [32-38] | [34] |
| TB notifications (per 100,000 population/year) | 2019 | 15-99 | 61.2 [49-73.5] | [34] | 2019 | 15-99 | 201.1 [160.9-241.4] | [34] |
| TB notifications (per 100,000 population/year) | 2019 | 0-99 | 50.8 [40.6-61] | [34] | 2019 | 0-99 | 158.2 [126.6-189.9] | [34] |
| TB incidence (per 100,000 population/year) | - | - | - | - | 2000 | 0-99 | 289 [99-578] | [35] |

| | | | | | | | | |
|---|---|---|---|---|---|---|---|---|
| TB incidence (per 100,000 population/year) | - | - | - | - | 2019 | 0-14 | 91.6 [55.8-127.6] | [35] |
| TB incidence (per 100,000 population/year) | - | - | - | - | 2019 | 15-99 | 229.4 [139.6-320.1] | [35] |
| TB incidence (per 100,000 population/year) | - | - | - | - | 2019 | 0-99 | 193.2 [125.9-259.8] | [35] |
| Mtb infection prevalence (proportion) | 2000 | 15-99 | 0.47 [0.42-0.51] | [36] | 2021 | 15-99 | 0.314 [0.114-0.514] | [32] |
| Protection against reinfection in early compartments | 2019 | 0-99 | 0.725 [0.6-0.85] | [20] | 2019 | 0-99 | 0.725 [0.6-0.85] | [20] |
| Reactivation rate in Infection-Slow Early | 2019 | 0-99 | 0.0043 [0.0020-0.0066] | [37] | 2019 | 0-99 | 0.0043 [0.0020-0.0066] | [37] |
| Reactivation rate in Infection-Slow Late | 2019 | 0-99 | 0.0006 [0.0002-0.001] | [37] | 2019 | 0-99 | 0.0006 [0.0002-0.001] | [37] |

*Implemented as 1-(proportion of overall population in the Uninfected compartment)

Table E. Targets used in the calibration process.

### Calibrated baseline scenarios for India

Figure C shows how different values of the self-clearance rate and of the natural protection against reinfection in self-cleared individuals affect the proportion of individuals in the various compartments and the distribution of the flows into the Infection-Fast compartment in India in 2025.

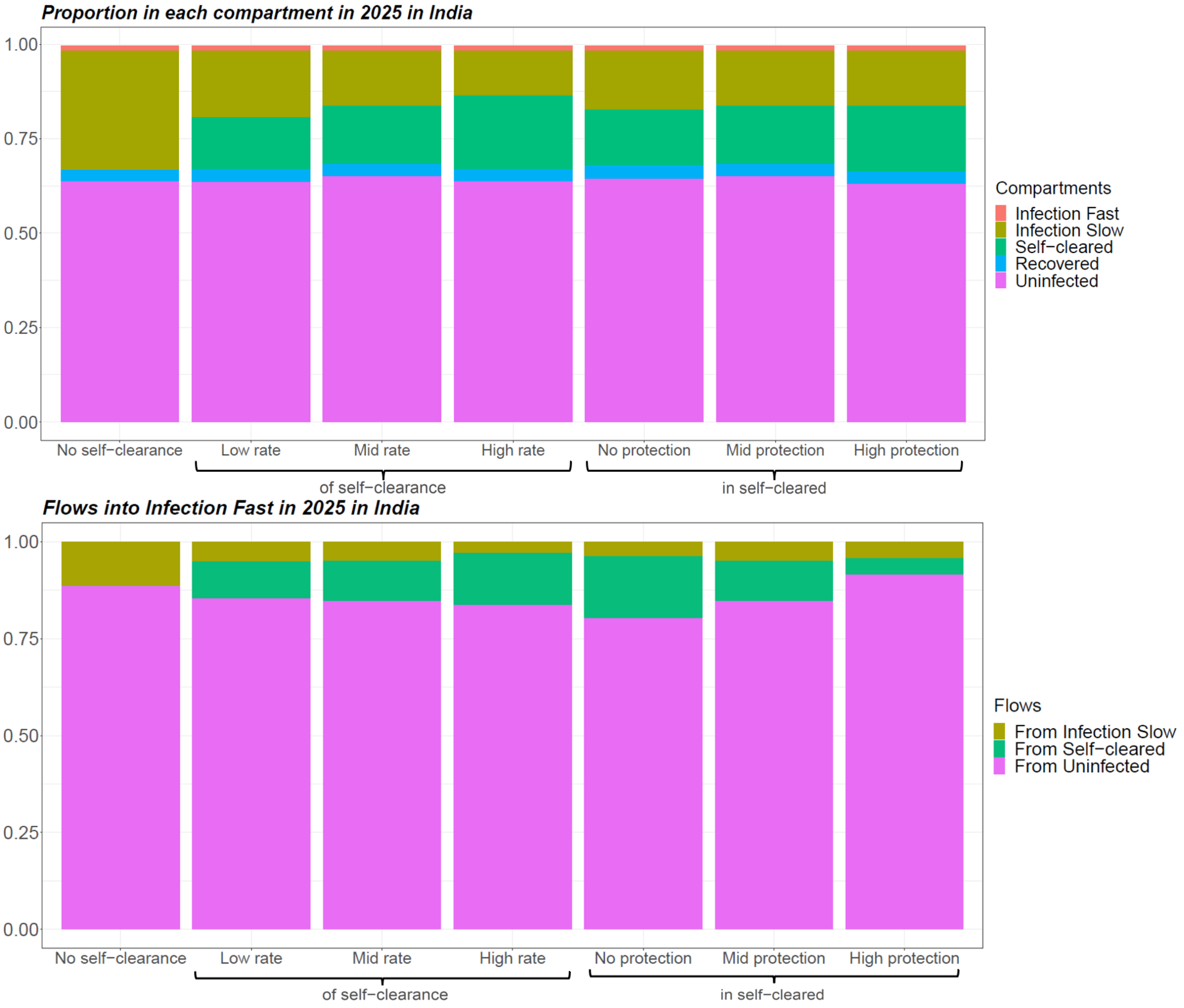


Figure C. Proportion of individuals in each compartment and distribution of flows into Infection-Fast in 2025 for different baseline scenarios in India. In scenarios with self-clearance, when the rate of self-clearance or the level of protection in Uninfected-Cleared individuals are not indicated, they are fixed at their mid value.

### C. Vaccine structure

Section C describes the vaccine structure based on Clark et al [8]. Before vaccination, all individuals in the model began in the Never Vaccinated compartment, with no vaccine protection. For the two “any infection” vaccines (cf. Figure D), upon vaccination, all individuals, apart from those in the Disease compartments or Resolved compartments, transitioned to the Vaccinated and Protected compartments, receiving the maximum (50%)

protection. Individuals in the Disease compartments or On-treatment compartment were not vaccinated. Since we are assuming lifelong protection from the vaccine, the only way of losing the protection once vaccinated was by entering the Disease Subclinical compartment: in such cases individuals transitioned to the Vaccinated and not Protected compartment.

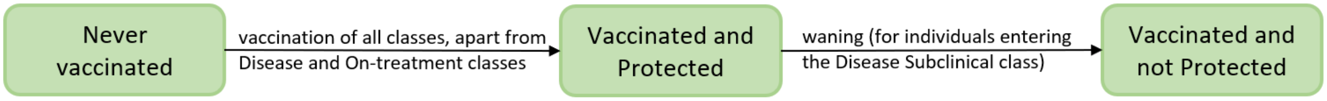


Figure D. Structure of “any infection vaccines”

For the no current infection vaccines and the current infection vaccines (cf. Figure E), upon vaccination, individuals in the Uninfected - Cleared compartments transitioned to the Vaccinated Cleared compartment, receiving 0%, 25%, 50% protection, depending on the vaccine scenario (see next section). All other individuals with the appropriate host infection status upon vaccination, transitioned instead to the Vaccinated and Protected compartment, receiving the maximum (50%) protection. As before, the only way of losing the protection once vaccinated was by developing tuberculosis disease.

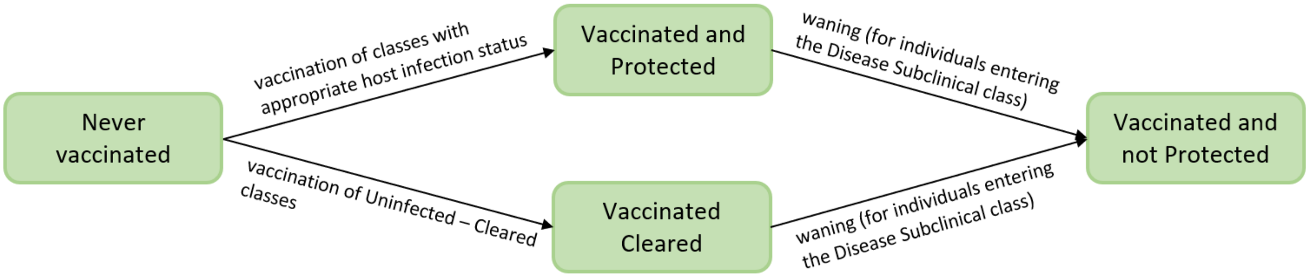


Figure E. Structure for no current infection vaccines and current infection vaccines.

### D. Maximum effect of self-clearance on vaccine impact

#### Maximum effect when the rate of self-clearance is within the range provided in [15]

Figure F shows the incidence reduction in 2050 for each no current infection and each current infection vaccine for all extreme scenarios, i.e. scenarios where the three self-clearance characteristics (self-clearance rate, level of protection against reinfection in self-cleared individuals and vaccine efficacy in self-cleared individuals) assumed either their minimum or maximum value.

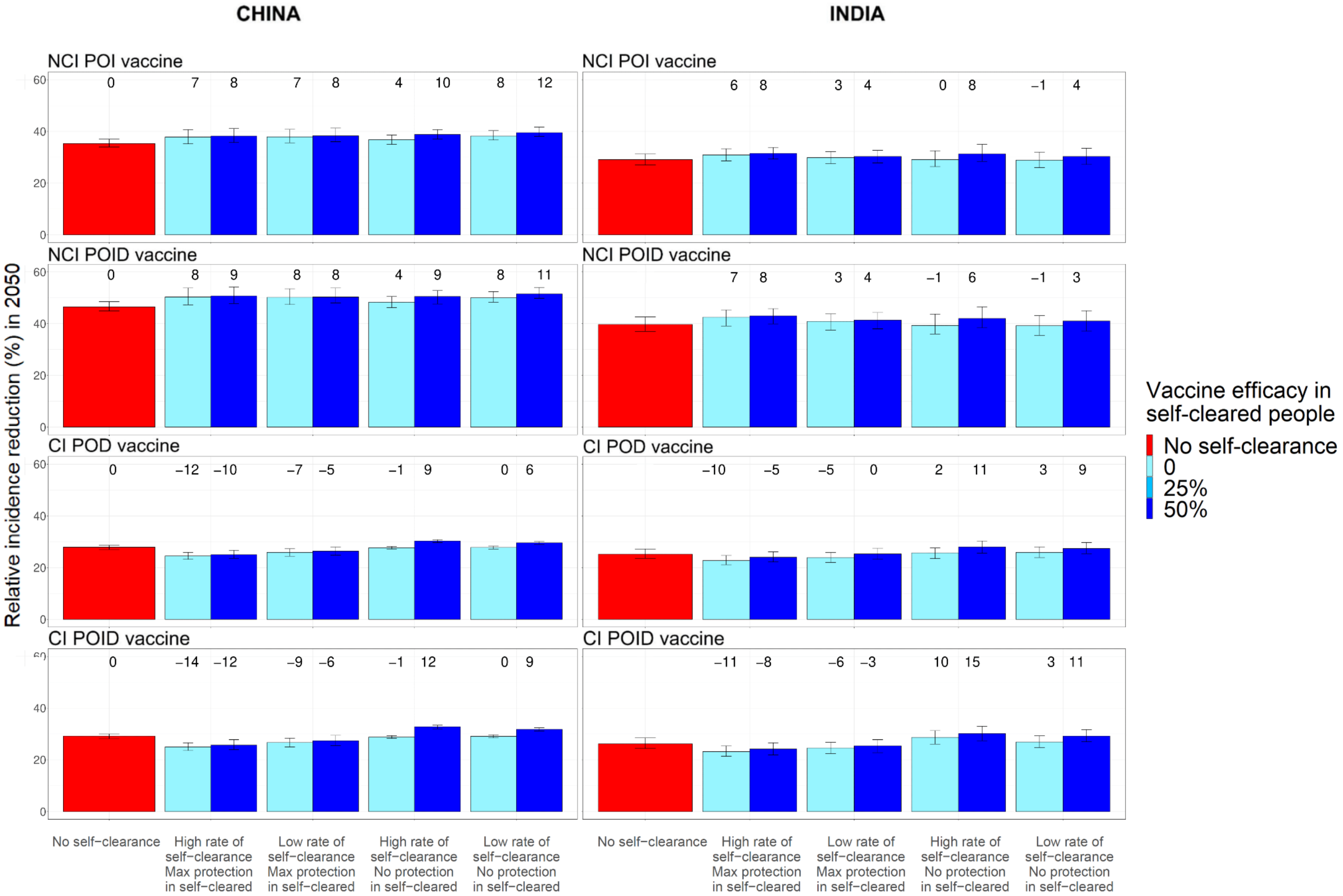


Figure F. Relative incidence reduction in 2050 compared to no self-clearance scenario, when varying all three self-clearance characteristics at once, for no current infection vaccines and current infection vaccines (China on the left, India on the right). The height of the columns show the median percentage reduction in incidence in 2050 compared to the scenario where no vaccine is implemented, with vertical bars indicating the 95% confidence interval. The numbers on top of each column correspond to the relative increase/decrease of the incidence reduction compared to the no self-clearance scenario. All extreme scenarios, i.e. scenarios where the three self-clearance characteristics (self-clearance rate, level of protection against reinfection in self-cleared individuals and vaccine efficacy in self-cleared individuals) assumed either their minimum or maximum value, are shown. NCI: no current infection, CI: current infection, POI: prevention of infection, POD: prevention of disease, POID: prevention of infection and disease.

#### Maximum effect when the rate of self-clearance is not within the range provided in [15]

We report below the analysis we conducted to estimate the maximum effect on vaccine impact that self-clearance may have if we allowed self-clearance rates higher than those found in [15]. Since for a fixed level of natural protection in self-cleared individuals, the sum of all individuals in the Infection Slow and Uninfected-Cleared compartments was constant for different self-clearance rates, we used this sum as an upper bound for the size of the Uninfected-Cleared compartments. For any given level of natural protection against reinfection in self-cleared individuals, we had three pairs (proportion of population in Uninfected-Cleared compartments, reduction in tuberculosis reduction in 2050 compared to the no-self-clearance scenario), obtained by setting the self-clearance rate to the 2.5 percentile, median or 97.5 percentile in [15]. We calculated the best-fitting straight line for the three available pairs and used it to estimate the effect on vaccine impact when the size of the Uninfected-Cleared compartments was equal to the calculated upper bound.

For each no current infection vaccine and current infection vaccine we varied the level of natural protection in self-cleared individuals (no protection and maximum protection) and the vaccine efficacy in self-cleared individuals (no efficacy, maximum efficacy). For each "any infection" vaccine we only varied the level of natural protection in self-cleared individuals (no protection and maximum protection), since "any infection" vaccines are assumed to take no matter the host infection status. As the only uncertainty is about the maximum self-clearance rate, combinations of level of natural protection and vaccine efficacy in self-cleared individuals for which the effect on vaccine impact reduced or did not substantially increase when the self-clearance rate increased are not shown below. In all cases reported below, the best fitting line had an $R^2$ of at least 0.9.

In Figure G we see an example of this process. It refers to China, current infection, prevention of infection and disease vaccine, when self-cleared individuals are assumed to have no natural protection against reinfection and the vaccine is assumed to take perfectly on self-cleared individuals. In this case the Uninfected-Cleared and Infection-Slow compartments accounted for 26% of the overall population in 2025, independently of the self-clearance rate used. The three blue points correspond to values obtained when setting the self-clearance rate to the lower bound, median and upper bound in [15], while the orange point gives the estimate of the effect on vaccine impact (+20%) in the extreme case where 26% of the population is in the Uninfected-Cleared compartments and no individual is in the Infection-Slow compartments.

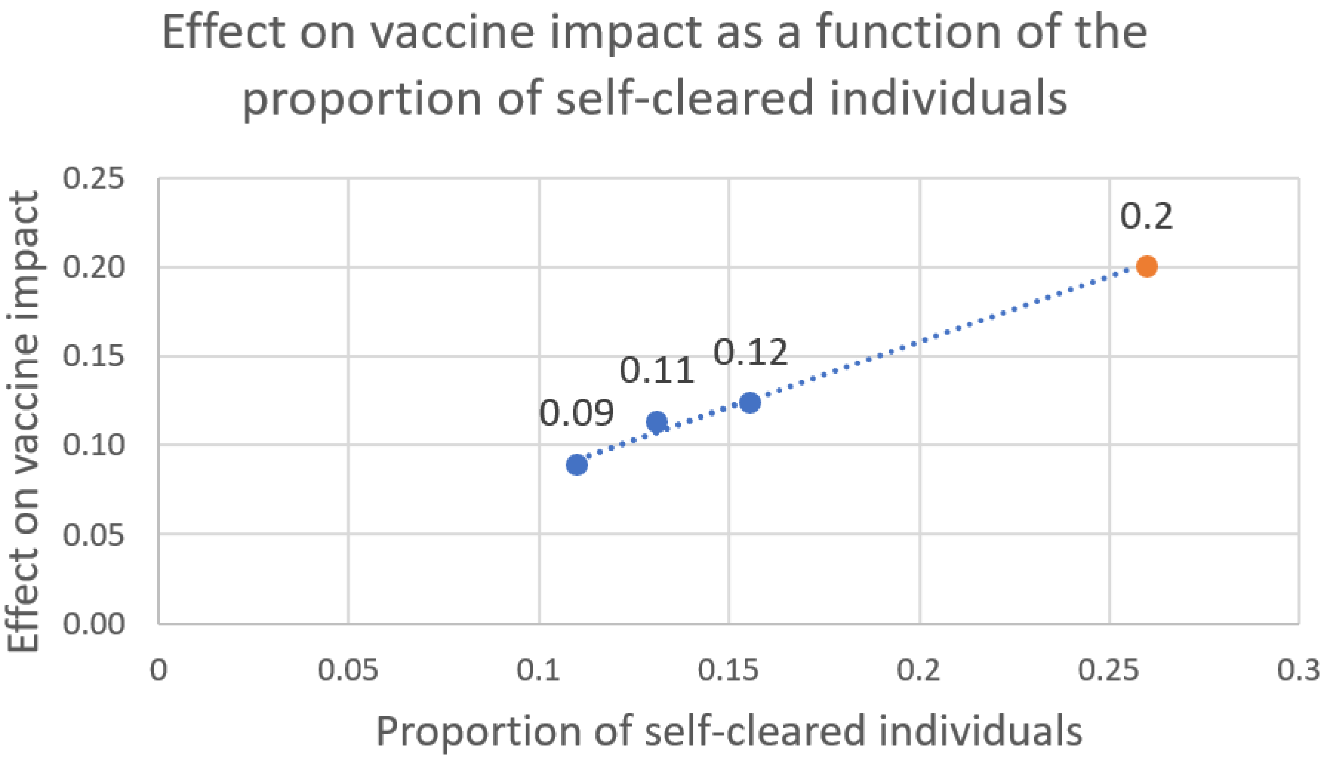


Figure G. Estimating the maximum possible effect on tuberculosis incidence reduction in 2050 when allowing self-clearance rates higher than those provided in [15]. The plot shows the analysis for the current infection, prevention of infection and disease vaccine in China, when self-cleared individuals are assumed to have no natural protection against reinfection and the vaccine is assumed to take perfectly on self-cleared individuals.

| China, CI POD, no natural protection and maximum vaccine efficacy in self-cleared individuals | |
|---|---|
| Proportion self-cleared | Effect on vaccine impact |
| 11% (obtained with low self-clearance rate) | +6% (from model) |
| 13% (obtained with mid self-clearance rate) | +7% (from model) |
| 16% (obtained with high self-clearance rate) | +9% (from model) |

| | |
|---|---|
| 26% (estimated assuming no individuals in Infection-Slow compartments) | +14% (estimated with best-fitting line) |

| China, CI POD, maximum natural protection and maximum vaccine efficacy in self-cleared individuals | |
|---|---|
| Proportion self-cleared | Effect on vaccine impact |
| 13% (obtained with low self-clearance rate) | -5% (from model) |
| 15% (obtained with mid self-clearance rate) | -9% (from model) |
| 18% (obtained with high self-clearance rate) | -10% (from model) |
| 28% (estimated assuming no individuals in Infection-Slow compartments) | -19% (estimated with best-fitting line) |

| China, CI POD, maximum natural protection and no vaccine efficacy in self-cleared individuals | |
|---|---|
| Proportion self-cleared | Effect on vaccine impact |
| 13% (obtained with low self-clearance rate) | -7% (from model) |
| 15% (obtained with mid self-clearance rate) | -10% (from model) |
| 18% (obtained with high self-clearance rate) | -12% (from model) |
| 28% (estimated assuming no individuals in Infection-Slow compartments) | -21% (estimated with best-fitting line) |

| China, CI POID, no natural protection and maximum vaccine efficacy in self-cleared individuals | |
|---|---|
| Proportion self-cleared | Effect on vaccine impact |
| 11% (obtained with low self-clearance rate) | +9% (from model) |
| 13% (obtained with mid self-clearance rate) | +11% (from model) |
| 16% (obtained with high self-clearance rate) | +12% (from model) |
| 26% (estimated assuming no individuals in Infection-Slow compartments) | +20% (estimated with best-fitting line) |

| China, CI POID, maximum natural protection and maximum vaccine efficacy in self-cleared individuals | |
|---|---|
| Proportion self-cleared | Effect on vaccine impact |

| | |
|---|---|
| 13% (obtained with low self-clearance rate) | -6% (from model) |
| 15% (obtained with mid self-clearance rate) | -7% (from model) |
| 18% (obtained with high self-clearance rate) | -12% (from model) |
| 28% (estimated assuming no individuals in Infection-Slow compartments) | -22% (estimated with best-fitting line) |

| China, CI POID, no natural protection and no vaccine efficacy in self-cleared individuals | |
|---|---|
| Proportion self-cleared | Effect on vaccine impact |
| 13% (obtained with low self-clearance rate) | -9% (from model) |
| 15% (obtained with mid self-clearance rate) | -12% (from model) |
| 18% (obtained with high self-clearance rate) | -14% (from model) |
| 28% (estimated assuming no individuals in Infection-Slow compartments) | -25% (estimated with best-fitting line) |

| India, NCI POI, no natural protection and maximum vaccine efficacy in self-cleared individuals | |
|---|---|
| Proportion self-cleared | Effect on vaccine impact |
| 13% (obtained with low self-clearance rate) | +4% (from model) |
| 15% (obtained with mid self-clearance rate) | +6 (from model) |
| 18% (obtained with high self-clearance rate) | +8% (from model) |
| 31% (estimated assuming no individuals in Infection-Slow compartments) | +16% (estimated with best-fitting line) |

| India, NCI POI, maximum natural protection and maximum vaccine efficacy in self-cleared individuals | |
|---|---|
| Proportion self-cleared | Effect on vaccine impact |
| 15% (obtained with low self-clearance rate) | +4% (from model) |
| 17% (obtained with mid self-clearance rate) | +6% (from model) |
| 21% (obtained with high self-clearance rate) | +8% (from model) |
| 32% (estimated assuming no individuals in Infection-Slow compartments) | +16% (estimated with best-fitting line) |

| India, NCI POI, maximum natural protection and no vaccine efficacy in self-cleared individuals | |
|---|---|
| Proportion self-cleared | Effect on vaccine impact |
| 15% (obtained with low self-clearance rate) | +3% (from model) |
| 17% (obtained with mid self-clearance rate) | +5% (from model) |
| 21% (obtained with high self-clearance rate) | +6% (from model) |
| 32% (estimated assuming no individuals in Infection-Slow compartments) | +13% (estimated with best-fitting line) |

| India, NCI POID, no natural protection and maximum vaccine efficacy in self-cleared individuals | |
|---|---|
| Proportion self-cleared | Effect on vaccine impact |
| 13% (obtained with low self-clearance rate) | +3% (from model) |
| 15% (obtained with mid self-clearance rate) | +4% (from model) |
| 18% (obtained with high self-clearance rate) | +6% (from model) |
| 31% (estimated assuming no individuals in Infection-Slow compartments) | +12% (estimated with best-fitting line) |

| India, NCI POID, maximum natural protection and maximum vaccine efficacy in self-cleared individuals | |
|---|---|
| Proportion self-cleared | Effect on vaccine impact |
| 15% (obtained with low self-clearance rate) | +4% (from model) |
| 17% (obtained with mid self-clearance rate) | +6% (from model) |
| 21% (obtained with high self-clearance rate) | +8% (from model) |
| 32% (estimated assuming no individuals in Infection-Slow compartments) | +16% (estimated with best-fitting line) |

| India, NCI POID, maximum natural protection and no vaccine efficacy in self-cleared individuals | |
|---|---|
| Proportion self-cleared | Effect on vaccine impact |
| 15% (obtained with low self-clearance rate) | +3% (from model) |
| 17% (obtained with mid self-clearance rate) | +6% (from model) |

| | |
|---|---|
| 21% (obtained with high self-clearance rate) | +7% (from model) |
| 32% (estimated assuming no individuals in Infection-Slow compartments) | +13% (estimated with best-fitting line) |

| India, CI POD, no natural protection and maximum vaccine efficacy in self-cleared individuals | |
|---|---|
| Proportion self-cleared | Effect on vaccine impact |
| 13% (obtained with low self-clearance rate) | +9% (from model) |
| 15% (obtained with mid self-clearance rate) | +10% (from model) |
| 18% (obtained with high self-clearance rate) | +11% (from model) |
| 31% (estimated assuming no individuals in Infection-Slow compartments) | +17% (estimated with best-fitting line) |

| India, NCI POID, maximum natural protection and no vaccine efficacy in self-cleared individuals | |
|---|---|
| Proportion self-cleared | Effect on vaccine impact |
| 15% (obtained with low self-clearance rate) | -5% (from model) |
| 17% (obtained with mid self-clearance rate) | -8% (from model) |
| 21% (obtained with high self-clearance rate) | -10% (from model) |
| 32% (estimated assuming no individuals in Infection-Slow compartments) | -19% (estimated with best-fitting line) |

| India, CI POID, no natural protection and maximum vaccine efficacy in self-cleared individuals | |
|---|---|
| Proportion self-cleared | Effect on vaccine impact |
| 13% (obtained with low self-clearance rate) | +11% (from model) |
| 15% (obtained with mid self-clearance rate) | +14% (from model) |
| 18% (obtained with high self-clearance rate) | +15% (from model) |
| 31% (estimated assuming no individuals in Infection-Slow compartments) | +25% (estimated with best-fitting line) |

| India, CI POID, maximum natural protection and maximum vaccine efficacy in self-cleared individuals |
|---|

| Proportion self-cleared | Effect on vaccine impact |
|---|---|
| 15% (obtained with low self-clearance rate) | -3% (from model) |
| 17% (obtained with mid self-clearance rate) | -4% (from model) |
| 21% (obtained with high self-clearance rate) | -8% (from model) |
| 32% (estimated assuming no individuals in Infection-Slow compartments) | -16% (estimated with best-fitting line) |

| India, CI POID, maximum natural protection and no vaccine efficacy in self-cleared individuals | |
|---|---|
| Proportion self-cleared | Effect on vaccine impact |
| 15% (obtained with low self-clearance rate) | -6% (from model) |
| 17% (obtained with mid self-clearance rate) | -9% (from model) |
| 21% (obtained with high self-clearance rate) | -11% (from model) |
| 32% (estimated assuming no individuals in Infection-Slow compartments) | -22% (estimated with best-fitting line) |

### E. Maximum effect of self-clearance for current infection vaccines

Figure H shows how the percentage reduction in tuberculosis incidence in 2050 varies for each “any infection” vaccine, when we vary the self-clearance rate and the level of natural protection in Uninfected-Cleared individuals at once (note that here we did not vary the vaccine efficacy in Uninfected-Cleared individuals, since “any infection” vaccines were assumed to take on all individuals, independently of their infection status). For each of these two characteristics, we explored the two extreme values, minimum and maximum, obtaining four possibilities for each vaccine. Here we see that self-clearance has very little effect on vaccine impact, with a maximum increase of 6% in China and 3% in India.

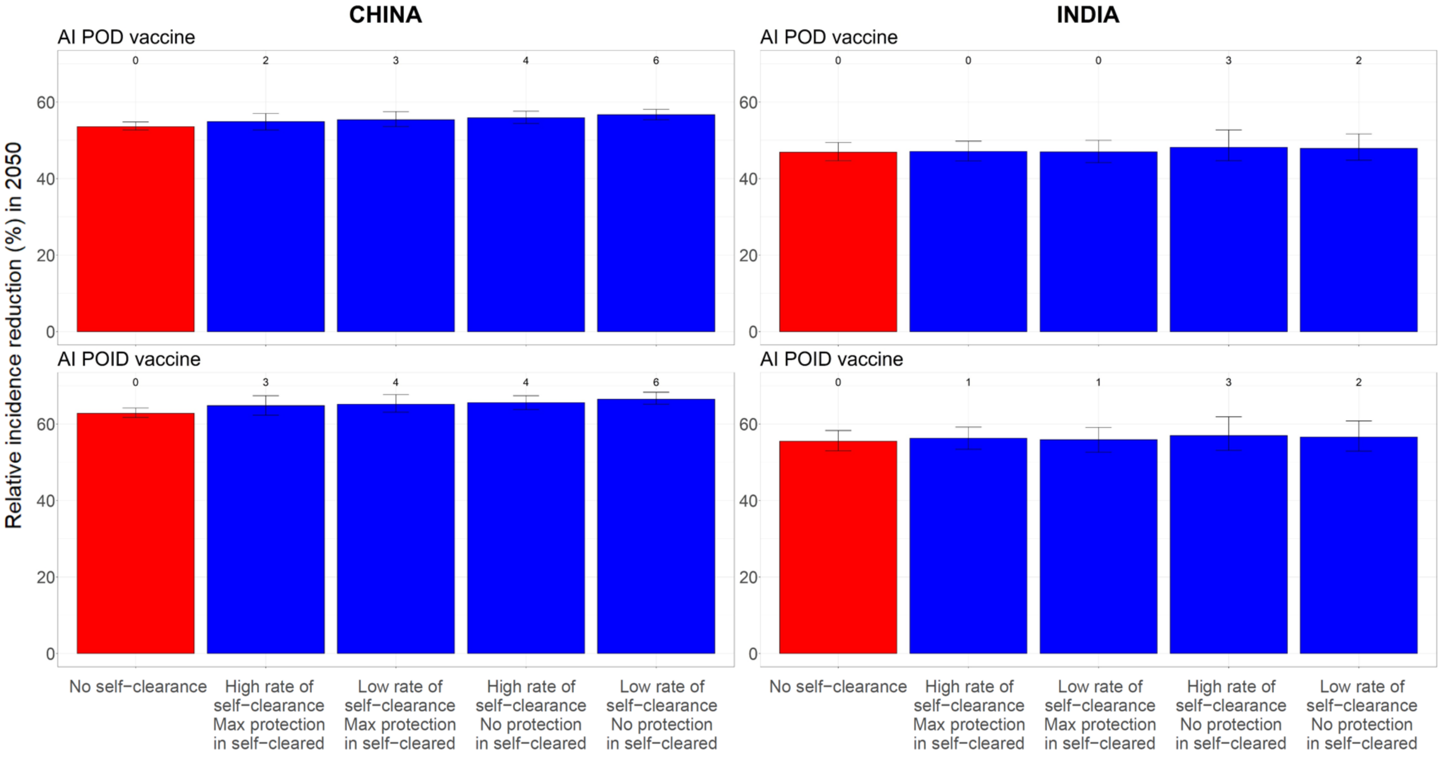


Figure H. Effect on vaccine impact when all self-clearance characteristics are varied at once for “any infection vaccines” (China on the left, India on the right). The height of the columns show the median percentage reduction in incidence in 2050 compared to the scenario where no vaccine is implemented, with vertical bars indicating the 95% confidence interval. The numbers on top of each column correspond to the relative increase/decrease of the incidence reduction compared to the no self-clearance scenario (red). Scenarios with self-clearance are in blue, while scenarios without self-clearance are in red. NCI: no current infection, CI: current infection, POI: prevention of infection, POD: prevention of disease, POID: prevention of infection and disease.
